# A Simplified Classification for Age-Related Macular Degeneration Based on Optical Coherence Tomography

**DOI:** 10.64898/2026.03.29.26349635

**Authors:** Tsai-Chu Yeh, Jonathan B. Lin, Prithvi Mruthyunjaya, Theodore Leng, Charles DeBoer, Yasir J. Sepah, David RP Almeida, Stephen Smith, Vinit B. Mahajan

## Abstract

**Background and Objective:** As optical coherence tomography (OCT) has enabled the identification of an expanding set of age-related macular degeneration (AMD) risk biomarkers and become central to routine clinical practice, there remains a need for a simplified grading scheme that allows physicians to communicate and synchronize AMD grading directly from standard OCT imaging rather than relying on traditional color fundus imaging. This study aims to establish a standardized OCT-based AMD classification that balances diagnostic accuracy with practicality for use across clinical and research settings.

**Patients and Methods:** Spectral-domain optical coherence tomography scans were independently graded by two retinal specialists following the newly proposed Stanford OCT-Based AMD Classification (SOAC). Discrepancies were adjudicated by a third independent retinal specialist. Intergrader agreement was assessed using weighted kappa coefficients.

**Results:** Among the 109 eyes from 108 patients (mean age 79.61□±□7.57 years; 41.7% male, 58.3% female), AMD staging based on SOAC was distributed as follows: normal aging in 9 patients (8.3%), early AMD in 16 (14.7%), intermediate AMD in 32 (29.4%), neovascular AMD (nAMD) in 18 (16.5%), geographic atrophy (GA) in 20 (18.3%), and combined nAMD and GA in 14 (12.8%). The overall intergrader agreement demonstrated robust consistency, with a weighted kappa value of 0.95 (95% CI: 0.92–0.98), signifying excellent intergrader reliability and reinforcing the validity of SOAC.

**Conclusion:** SOAC provides a standardized, OCT-based framework for AMD grading that demonstrates high intergrader agreement. By enabling consistent classification from commonly acquired OCT scans, SOAC supports reliable disease staging and facilitates integration across clinical studies and translational research. As imaging and molecular data continue to expand, SOAC can serve as a common OCT-based reference for phenotype refinement and longitudinal AMD studies.

**Key questions:** **What is already known on this topic?**

- Existing classifications of age-related macular degeneration (AMD) rely heavily on color fundus photography and inconsistently incorporate optical coherence tomography (OCT) biomarkers, despite OCT’s superior resolution for detecting subclinical structural changes.

**What this study adds?**

- This study introduces an OCT-based classification framework that consolidates key biomarkers with proven prognostic relevance into a simplified, tiered staging protocol. By prioritizing a curated set of evidence-supported, high-risk OCT biomarkers, the Stanford OCT-Based AMD Classification (SOAC) provides a common framework and shared language for OCT-based AMD staging, facilitates longitudinal monitoring, and reduces intergrader variability through standardized reporting guidelines. The framework deliberately balances clinical practicality with scientific rigor, excluding less significant or potentially confusing features to ensure scalability across diverse clinical and research settings.

**How this study might affect research, practice or policy?**

- SOAC provides an OCT-based framework for AMD staging with high inter-physician agreement. This is particularly important in real-world clinical and translational research settings, where color fundus photography is not always available. By offering a common OCT-based reference, SOAC reduces diagnostic variability and supports more consistent AMD staging across clinicians and studies. The classification incorporates up-to-date, evidence-based, high-risk OCT features associated with disease progression and is designed to be scalable, allowing integration with emerging multimodal data, including AI-driven imaging analysis and molecular profiling. By bridging advances in retinal imaging with practical clinical application, SOAC lays the groundwork for improved risk stratification, longitudinal outcome studies, and more standardized reporting in AMD.

## Introduction

Age-related macular degeneration (AMD) is a leading cause of vision loss in older adults worldwide, profoundly impacting patient quality of life and placing significant burdens on healthcare systems.^1,2^ Characterized by progressive macular degeneration, AMD manifests across a clinical spectrum from early-stage drusen accumulation to late-stage geographic atrophy (GA) and macular neovascularization (MNV) in neovascular AMD (nAMD).^3,4^ Accurate and consistent staging of AMD is critical for optimizing therapeutic interventions, predicting trajectories of visual outcome, and tailoring individualized management strategies.

Optical coherence tomography (OCT) has revolutionized the diagnosis and management of AMD by enabling noninvasive visualization of retinal microstructure. Over the past decade, numerous OCT biomarkers have been described, including retinal and choroidal lesions, morphological features, and reflectivity patterns.^5,6^ Recent consensus efforts have sought to standardize these biomarkers to facilitate their use in clinical trials.^7^ Nevertheless, current AMD grading in routine clinical practice remains largely based on funduscopic findings rather than OCT-derived structural features.^8^ Although some studies have attempted to translate traditional color fundus-based grading schemes into OCT-based classifications for specific research purposes^9^, these efforts have not evaluated inter-physician agreement or grading consistency. Importantly, the need for an OCT-based grading framework is further reinforced by contemporary reimbursement structures, in which OCT has increasingly replaced color fundus photography as the primary imaging modality in AMD care.

To address this gap, we developed the Stanford OCT-Based AMD Classification (SOAC), an OCT-centered framework that consolidates biomarkers with established clinical and prognostic relevance into a simplified, practical staging system. SOAC prioritizes OCT features that have shown strong correlations with disease progression and outcomes, providing a structured approach for consistent grading, risk stratification, and longitudinal disease assessment. SOAC is not designed to replace traditional color fundus photograph-based classifications, but to complement and align with them, facilitating a smoother transition toward OCT as a primary imaging modality. SOAC incorporates several deliberate design choices informed by emerging clinical evidence. It includes a category with concurrent nAMD and GA, reflecting increasing recognition that the high coexistence rate represents a distinct clinical phenotype. In addition, SOAC emphasizes drusen burden and extent rather than drusen size alone, aligning with AREDS-based frameworks.^10^ To support real-world applicability, we assessed intergrader agreement using SOAC and observed consistent grading among readers. While future longitudinal studies will be needed to directly compare the prognostic performance of SOAC with existing classification systems, this initial evaluation establishes a reproducible framework suitable for further validation.

## Methods

### Study Design and Population

This retrospective study included patients aged ≥55 years with varying degrees of AMD. Patients were consecutively selected from a clinical database between October 2021 and January 2024, with inclusion criteria requiring a diagnosis of AMD in at least one eye, as documented in the electronic health record. Exclusion criteria included eyes with other macular pathologies from non-AMD conditions (e.g., diabetic macular edema, retinal vein occlusion, myopic maculopathy). This study was approved by the XXX Institutional Review Board and adhered to the principles of the Declaration of Helsinki.

### Imaging Protocol

Macular imaging was performed using the Cirrus HD-OCT 5000 system (Carl Zeiss Meditec, Inc., Dublin, CA, USA, v.11.5), utilizing the Macular Cube 512×128 scan protocol. This standardized 6 × 6 mm volumetric scan was centered on the fovea, capturing 128 horizontal B-scans, each consisting of 512 A-scans per line, enabling high-definition visualization of retinal layers and subretinal structures across the central macula.

### Imaging Assessment

OCT scans were systematically analyzed and graded by two independent, board-certified retinal specialists in accordance with the newly proposed Stanford OCT-Based AMD Classification (SOAC). Intergrader discrepancies in AMD classification were adjudicated by a third retinal specialist. The primary outcome was intergrader agreement, quantified using the weighted kappa coefficient. Graders were blinded to clinical data and restricted to OCT scans only during initial grading. For adjudication, longitudinal clinical history were incorporated as needed to reconcile discordant classifications.

### Grading Criteria and Measurement Protocol

SOAC was based on the number and size of drusen, presence of reticular pseudodrusen (RPD), intraretinal hyperreflective foci (HRF), abnormalities in the external limiting membrane (ELM), ellipsoid zone (EZ), or interdigitation zone (IZ), as well as the presence of MNV, hemorrhage, exudation, subretinal fluid (SRF), intraretinal fluid (IRF), fibrosis, and retinal pigment epithelium (RPE) and outer retinal atrophy, which are indicative of late stage AMD. Drusen classification was based on size using the built-in caliper tool of the Zeiss FORUM viewer (v4.4), with small drusen defined as <63 µm, medium drusen as 63–124 µm, and large drusen as ≥125 µm. Patients were classified into six AMD categories **(Figure 1)**:

**Figure 1.**
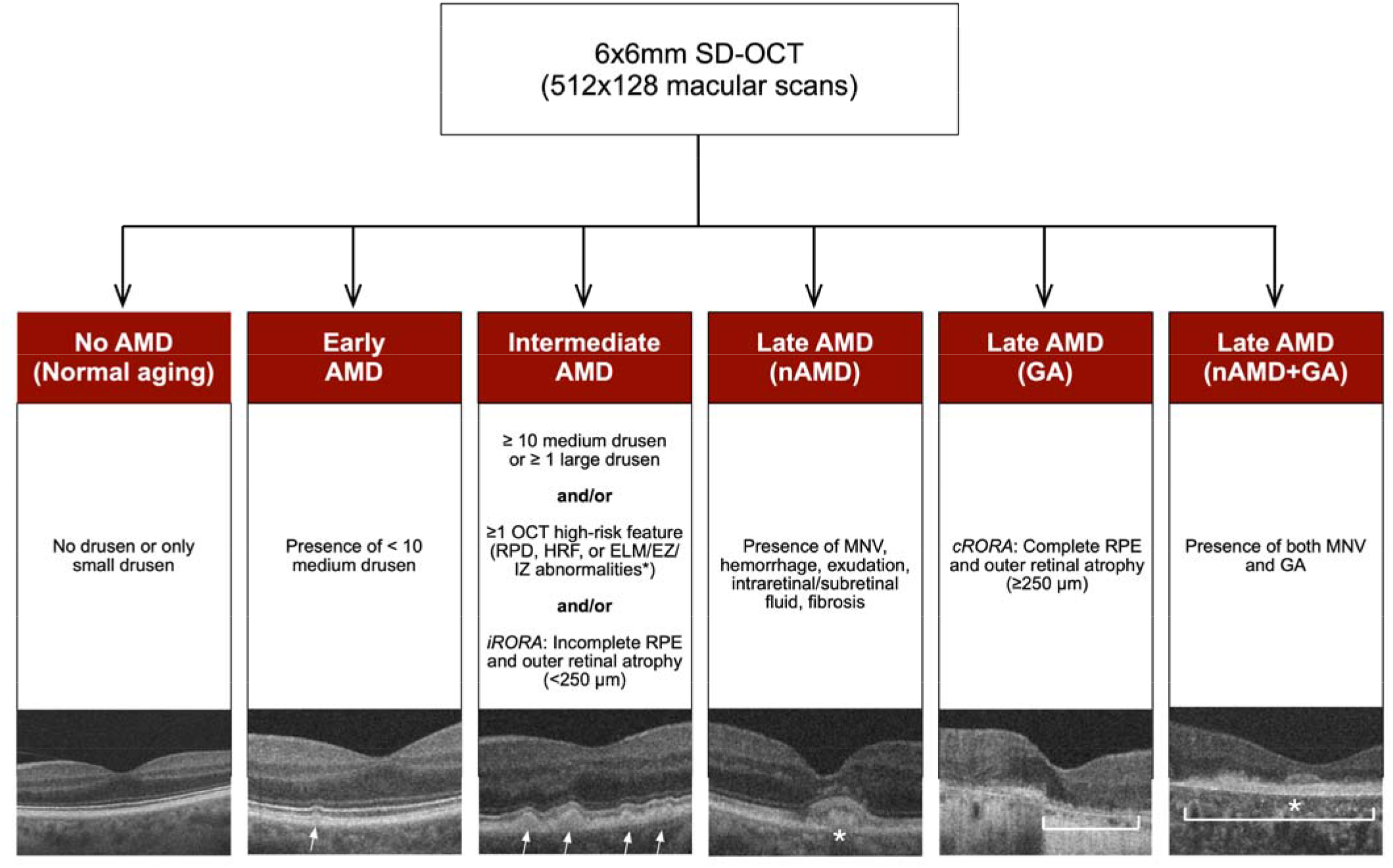
Stanford OCT-Based AMD Classification (SOAC). 6×6 mm Spectral-domain optical coherence tomography (SD-OCT) macular volumes (512 × 128 scans) were evaluated to classify age-related macular degeneration (AMD) based on OCT-defined structural biomarkers. No AMD (normal aging) was defined by the absence of drusen or the presence of only small drusen (<63 µm). Early AMD was defined by the presence of fewer than 10 medium drusen (63–124 µm) without OCT high-risk features. Intermediate AMD was defined by the presence of ≥10 medium drusen (63–124 µm) or ≥1 large drusen (≥125 µm), and/or the presence of at least one OCT high-risk feature, including reticular pseudodrusen (RPD), intraretinal hyperreflective foci (HRF), abnormalities of the external limiting membrane (ELM), ellipsoid zone (EZ), or interdigitation zone (IZ), and/or incomplete RPE and outer retinal atrophy (iRORA; <250 µm).Late AMD includes neovascular AMD (nAMD), defined by macular neovascularization (MNV; asterisk) with associated hemorrhage, exudation, intraretinal or subretinal fluid, or fibrosis; geographic atrophy (GA), defined by complete RPE and outer retinal atrophy (cRORA) with photoreceptor loss and hypertransmission (≥250 µm; bracket); or combined nAMD and GA. Representative B-scans are shown for each category. *ELM/EZ/IZ abnormalities were defined as any irregularity, attenuation, or disruption of the respective outer retinal layers.

#### No AMD (Normal aging)

No drusen or presence of only small drusen (<63 µm).

#### Early AMD

Presence of fewer than ten medium drusen (63–124 µm) without OCT high-risk features.

#### Intermediate AMD

Ten or more medium drusen or at least one large drusen, and/or presence of ≥1 OCT high-risk feature, including RPD, HRF or abnormalities of the ELM/EZ/IZ (irregularity, attenuation, or disruption), and/or incomplete RPE and outer retinal atrophy (iRORA, <250 µm).

#### Late AMD (nAMD)

Presence of MNV, with associated hemorrhage, exudation, intraretinal or subretinal fluid, or fibrosis.

#### Late AMD (GA)

Complete RPE and outer retinal atrophy (cRORA), defined as a well-demarcated area ≥250 µm characterized by complete RPE loss, overlying photoreceptor loss, and associated choroidal hypertransmission, consistent with CAM criteria.^11^

#### Late AMD (nAMD + GA)

Presence of both MNV and GA in the same eye.

### Statistical Analysis

Intergrader agreement was assessed using quadratic-weighted Cohen’s kappa statistics. Descriptive statistics were used to summarize the baseline demographics and distribution of AMD stages.

## Results

Among the 109 eyes from 108 patients included in the study, 41.7% were male (n = 45) and 58.3% were female (n = 63). The mean age was 79.61□±□7.57 years. The SOAC staging with adjudication consensus for AMD across the cohort is as follows **(Table 1)**: normal aging in 9 participants (8.3%), early AMD in 16 participants (14.7%), intermediate AMD in 32 participants (29.4%), nAMD in 18 participants (16.5%), GA in 20 participants (18.3%), and both nAMD and GA in 14 participants (12.8%).

**Table 1.** Patient Demographics for the Study Cohort.

|  | n (%) |
| --- | --- |
| **Total Participants** | 108 |
| **Male** | 45 (41.7%) |
| **Female** | 63 (58.3%) |
| **Age (Mean ± SD)** | 79.61 ± 7.57 |
| **SOAC AMD Staging (per eye)** |  |
| **Normal Aging** | 9 (8.3%) |
| **Early AMD** | 16 (14.7%) |
| **Intermediate AMD** | 32 (29.4%) |
| **Late AMD - nAMD** | 18 (16.5%) |
| **Late AMD - GA** | 20 (18.3%) |
| **Late AMD - nAMD & GA** | 14 (12.8%) |

\* SOAC: Stanford OCT-Based AMD Classification; nAMD: Neovascular age-related macular degeneration; GA: Geographic atrophy

### Intergrader Agreement

Intergrader agreement was assessed using Kappa statistics (κ) and demonstrated an overall robust consistency (unweighted Kappa = 0.85, 95% CI: 0.77–0.92; Quadratic-weighted Kappa = 0.95, 95% CI: 0.92– 0.98). This high level of intergrader reliability highlights the reproducibility of clinical phenotypic assessment and staging using this new grading scheme. A detailed breakdown of intergrader agreement by AMD classification is as follows **(Table 2)**: Normal aging had a kappa of 0.78 (95% CI: 0.57–0.99), while early AMD showed a kappa of 0.77 (95% CI: 0.59–0.95). Intermediate AMD had the highest agreement (κ = 0.91, 95% CI: 0.82–1.00). The kappa values for nAMD and GA were 0.90 (95% CI: 0.79–1.00) and 0.87 (95% CI: 0.74–1.00), respectively. Cases classified as both nAMD and GA had a kappa of 0.82 (95% CI: 0.67–0.97).

**Table 2.**
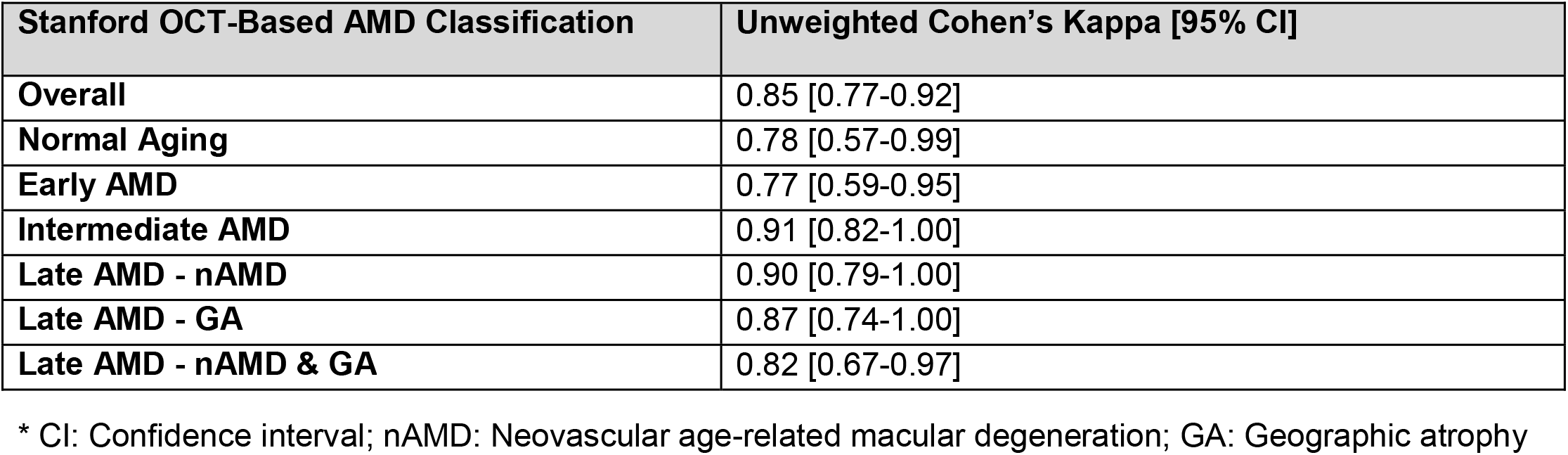
Intergrader Agreement in AMD Subgroup Classification.

## Discussion

OCT has revolutionized AMD evaluation, revealing structural biomarkers that were previously inaccessible with conventional imaging. However, the lack of consensus on which OCT biomarkers should be prioritized for grading AMD stages remains a significant challenge. While features such as drusen morphology, intraretinal HRF, exudates, and outer retinal atrophy are well-documented,^11-15^ their relative clinical significance and weighting in disease progression remain contentious, complicating efforts to establish a simplified and unified classification system. Additionally, existing classification frameworks have historically relied on color fundus-based systems like the Beckman classification^8^ and AREDS/AREDS2,^16,17^ which set the foundation for disease staging but do not fully capture the depth-resolved changes seen on OCT. There have been studies exploring OCT-based classification systems, with some incorporating artificial intelligence and deep learning.^18–20^ However, these approaches rely heavily on specific algorithms, software tools, or platforms, which may hinder their universal adoption, especially in resource-diverse settings. Overcoming these barriers is essential for developing a widely accepted OCT-driven classification system for AMD and ensuring its seamless integration into existing clinical workflows.

To address this gap, we propose SOAC, which incorporates established epidemiologic insights with validated high-risk OCT biomarkers to create a comprehensive and prognostically relevant framework. Designed for seamless integration across different clinical settings, SOAC aims to bridge the gap between traditional fundus-based classifications and recent OCT-driven evidence, offering a standardized yet adaptable classification for both clinical and research purposes. SOAC integrates foundational AMD classification principles from landmark population-based studies such as the Beaver Dam Eye Study (BDES),^21^ AREDS/AREDS2,^16,17^ the Blue Mountains Eye Study (BMES),^22^ and the Rotterdam Study.^23^ We incorporate drusen burden and extent, rather than relying only on drusen size thresholds, drawing on the AREDS/AREDS2 concept of drusen area from color fundus photography and adapting it to an OCT-based framework. Importantly, SOAC also incorporates a key principle from the Beckman classification by defining a “normal aging” category, which aims to distinguish benign age-related retinal changes from early AMD.^8^ This distinction is critical for ensuring accurate diagnosis and avoiding overdiagnosis.

In addition to drawing on landmark clinical trials and epidemiologic studies, SOAC incorporates more recent evidence from OCT-based biomarker research. RPD has been increasingly recognized as an important high-risk OCT feature associated with AMD progression. Our approach was guided by updated AREDS severity scales, which identify RPD as a significant risk factor and consider its presence as a binary feature for risk stratification.^24^ A recent meta-analysis by Trinh et al. identified 114 quantified OCT prognostic biomarkers and highlighted those most strongly associated with progression to late AMD, including ELM/EZ/IZ abnormalities, large drusen, RPD, and intraretinal HRF.^5^ Together, these updated insights informed our feature selection process, through which we curated a focused set of OCT imaging features with the most consistent and robust prognostic evidence. We also applied the latest CAM consensus definitions for RPE and outer retinal atrophy^15^, distinguishing iRORA from cRORA, with GA defined by the presence of cRORA in accordance with current clinical trial standards. While other AMD subgroups exist, including MNV subtypes and some OCT biomarkers in nAMD^25-27^, SOAC intentionally balances granularity with simplicity. This approach enables consistent application across clinical practice and research settings, supports standardized communication among clinicians and investigators, and remains adaptable as emerging imaging biomarkers and AI-based methods continue to evolve.

SOAC also demonstrated high intergrader agreement providing a critical validation of its reliability in real-world practice. This consistency reinforces its potential as a pragmatic standard for universal adoption. Intergrader discrepancies in this study primarily arose from challenges in distinguishing small drusen from normal aging changes, particularly in cases with hazy media or poor image quality. Additionally, differentiating cystoid degeneration from GA versus true IRF from nAMD posed another source of diagnostic discrepancy, especially when relying on a single OCT scan. However, upon closer review, we found that these variations could often be resolved through longitudinal OCT scans and by integrating clinical history, which provided additional context to refine classification accuracy.

While SOAC is grounded in clinical phenotyping, it was initially developed to support translational studies, including omics analyses, in settings where color fundus photography is not consistently available. By relying on OCT-defined structural features that are routinely acquired in modern practice, SOAC provides a practical and scalable framework for linking retinal phenotypes with molecular signatures. As genomic, proteomic, and other molecular profiling approaches increasingly shape AMD research and therapeutic development, an OCT-based system such as SOAC offers a common clinical reference for integrating imaging with biological data. Ultimately, SOAC is designed to support a shift from purely morphology-based staging toward a more molecularly informed classification of AMD, enabling improved risk stratification, patient selection, and precision therapeutics.

One limitation of this study reflects the shift in clinical practice away from color fundus photography as the primary imaging modality for AMD diagnosis and classification. While fundus photography has historically played a central role, it has largely been replaced by OCT in both routine clinical care and AMD research, largely due to reimbursement considerations and OCT’s superior structural resolution and depth-resolved imaging. As a result, direct comparisons with older fundus-based criteria are challenging. However, we took an additional step by reviewing AMD grading as documented in clinical charts, which likely reflects prior consensus staging frameworks. Overall agreement was high, particularly for nAMD and GA. Nevertheless, a substantial proportion of charts, mostly those corresponding to normal aging, early AMD, and intermediate AMD, did not include formal AMD staging. Instead, findings such as “drusen” or “AMD” were documented descriptively without explicit assignment of a disease stage. Future prospective and longitudinal studies that incorporate both fundus-based and OCT-based grading systems will be important for assessing their relationship to risk stratification and clinical outcomes. Another limitation is the difference in imaging coverage between OCT and color fundus photography, which complicates direct translation of fundus-based AMD classifications to OCT. In AREDS and AREDS2, grading was performed on stereoscopic color fundus photographs evaluated within a 6-mm diameter circular grading zone (the AREDS grid) centered on the fovea. In contrast, standard macular OCT volumes typically sample a 6×6 mm square area, with different geometry and sampling characteristics. This mismatch in spatial coverage makes one-to-one application of color fundus-based drusen criteria to OCT inherently challenging, particularly for assessing drusen extent in early and intermediate AMD. Finally, the relatively small sample size of this pilot study limits definitive conclusions regarding disease progression. Ongoing prospective cohort studies and planned multicenter validation will expand sample size, enable longitudinal assessment of progression to late AMD, and support further validation of SOAC.

In conclusion, SOAC introduces a fully OCT-based AMD grading system that enables disease staging without reliance on color fundus photography, reflecting the ongoing shift in real-world clinical practice. By integrating well-validated high-risk OCT biomarkers with updated concepts of OCT-defined atrophy, SOAC provides a clinically grounded yet forward-looking framework. The high intergrader agreement observed in this study supports its reproducibility and feasibility for routine use. Beyond clinical classification, SOAC offers a standardized structural phenotype well suited for translational research, including omics-based studies and emerging AI-driven imaging analyses, serving as a practical bridge between retinal imaging and biologically informed AMD stratification.

## Data Availability

All data produced in the present study are available upon reasonable request to the authors

## Institutional Review Board Statement

The study was approved by the Stanford University Institutional Review Board (IRB 39760) and adhered to the tenets set forth in the Declaration of Helsinki.

## Data Availability Statement

Not applicable.

